# Low Birth Weight Neonates in The Gambia & Burkina Faso: Prevalence, risk factors, and short-term adverse outcomes from a prospective observational study utilising trial data

**DOI:** 10.1101/2025.10.28.25338453

**Authors:** Bully Camara, Christian Bottomley, Athanase M. Some, Nathalie Beloum, Fatoumata Sillah, Joquina Chiquita Jones, Madikoi Danso, Usman N Nakakana, Shashu Graves, Sawadogo Yabre Edmond, Nassa Guetawende Job Wilfried, Joel D Bognini, Toussaint Rouamba, Marc C Tahita, Abdoulie Suso, Sulayman Bah, Omar B. Jarra, Siaka Badjie, Edrissa Sabally, Fatoumata Dibba, Kebba Manneh, Yusupha Njie, Hien So-vii Franck, Ebrahim Ndure, Umberto D’Alessandro, Halidou Tinto, Helen Brotherton, Anna Roca

**Affiliations:** Medical Research Council Unit The Gambia at London School of Hygiene and Tropical Medicine (MRCG at LSHTM), Fajara, The Gambia; London School of Hygiene and Tropical Medicine (LSHTM), London, United Kingdom; Institut de Recherche en Sciences de la Sante-Unité de recherche Clinique de Nanoro (IRSS-CRUN), Nanoro, Burkina Faso; Bill and Melinda Gates Foundation, Seattle, United States; Sherwood Forest NHS Trust, United Kingdom; Centre de Recherche en Epidémiologie, Biostatistique Et Recherche Clinique, Ecole de Santé Publique, Université Libre de Bruxelles, Bruxelles, Belgium; Ministry of Health, Banjul, The Gambia; Bundung Maternal and Child Health Hospital (BMCHH), Bundung, The Gambia; Victoria Hospital Kirkcaldy, NHS Fife, UK; ISGlobal, Campus Hospital Clínic-Universitat de Barcelona, Barcelona, Spain; ICREA, Pg. Lluís Companys 23, Barcelona, Spain

**Keywords:** Low birth weight, prevalence, risk factors, perinatal and neonatal, adverse outcomes, The Gambia, Burkina Faso

## Abstract

**Introduction:** Approximately 20 million neonates are born with low birth weight (LBW) annually, with West Africa bearing the highest burden of LBW and associated poor outcomes. This study aims to estimate the prevalence of LBW, identify maternal and foetal risk factors, and describe associated adverse perinatal and neonatal outcomes in a West African cohort.

**Methods:** These are secondary analyses of data from a randomised clinical trial (PregnAnZI-2). Pregnant women without known acute or chronic conditions were enrolled during labour at ten primary and secondary health facilities in The Gambia and Burkina Faso (2017–2021). Birth weight was measured within 24h of birth using standardised methods. Logistic regression was applied to identify associations between LBW and both selected risk factors and adverse outcomes, guided by a novel conceptual framework.

**Results:** 11,980 women and their 12,027 offspring were included in this study. The overall prevalence of LBW was 9.6% (1159/12027): 8.2% (551/6681) in The Gambia and 11.4% (608/5346) in Burkina Faso. Maternal risk factors for LBW included maternal ethnicity (*p*<0.001), primiparity (aOR 1.63, 9% CI 1.33-1.99, *p*<0.001), and previous history of stillbirth (aOR 1.88, 95% CI 1.35-2.62, *p*<0.001), while foetal risk factors included female sex (aOR 1.58, 95% CI 1.38-1.80, *p*<0.001) and twin birth (aOR 22.96, 95% CI 18.51-28.48, *p*<0.001). LBW newborns had increased risk of neonatal mortality (cOR 4.35, 95% CI 3.25-5.81, p<0.001); intrapartum stillbirth (cOR 3.37, 95% CI 2.07-5.51, p<0.001); neonatal hospitalisation (cOR 2.17, 95% CI 1.70-2.77, p<0.001); and intrapartum-related asphyxia defined as 1-min Apgar score <7 (cOR 2.04, 95% CI 1.59-2.62, p<0.001). 30.7% (69/225) of neonatal deaths and 26.2% (22/84) of intrapartum stillbirths were attributable to LBW.

**Conclusion:** This study underlines the substantial contribution of LBW towards adverse perinatal and neonatal outcomes in West Africa. Early identification of in-utero growth restriction and women at risk of preterm delivery could enable targeted antenatal interventions and timely referral for hospital delivery, improving perinatal and neonatal outcomes to reach Sustainable Development Goal 3.2.

## INTRODUCTION

Low birth weight (LBW), defined as a birth weight below 2.5 kg, remains a major contributor to neonatal mortality, accounting for 60% to 80% of neonatal deaths globally (1). It is also strongly associated with an increased risk of stillbirth (2). In 2020, an estimated 19.8 million infants were born with LBW, representing 1 in 7 (15%) newborns worldwide, with over 70% of these births occurring in low- and middle-income countries (LMICs) (3). Beyond early death, LBW infants face higher risks of long-term health challenges, including stunted growth (4), lower cognitive development (5), obesity, cardiovascular disease, and diabetes(6), compared to babies born with a normal birth weight (7). The consequences extend beyond individual health, but lead to substantial losses in human capital and long-term individual and societal impact (8).

Birth weight is influenced by a complex interplay of factors affecting foetal growth and pregnancy duration (9). Maternal undernutrition, infections, placental disorders, obstetric complications such as hypertensive disorders of pregnancy, and harmful environmental exposures can all interfere with foetal growth and development, resulting in LBW (9). LBW may arise from foetal growth restriction (FGR), preterm birth (PTB), or a combination of both (10). While FGR and PTB are predominant causes of LBW in LMICs (11) PTB-LBW is more prevalent in high-income countries, in part due to an increased frequency of multiple preterm births associated with assisted reproductive technologies (12).

Over the past two decades, global efforts to reduce LBW have intensified(13). In 2012, a global commitment was made to reduce LBW prevalence from 15% to 10.5% by 2025 (a 30% relative reduction) (14). However, progress has been slower than anticipated (15). Improving birthweight data coverage and quality is crucial to accelerating such progress. While more than 80% of births worldwide occur in health facilities, most data on LBW from high-burden regions is still derived from household surveys, which are prone to bias due to missing and inaccurate birthweight data (16). The sickest and smallest newborns, including those who die shortly after birth, are more likely to be excluded from these datasets (16).

In this study, we aimed to assess the prevalence of LBW and to examine associated risk factors and adverse perinatal and neonatal outcomes among women delivering at health facilities in The Gambia and Burkina Faso, including stillbirths. By addressing a critical data gap in West African countries, this study provides insights to the contribution of LBW to perinatal and neonatal morbidity and mortality.

## METHODS

### Study design

This study comprised of secondary data analyses from the PregnAnZI-2 trial (clinicaltrials.gov ref: NCT03199547); a phase III, double-blind, placebo-controlled randomised clinical trial in which ∼12000 women in The Gambia and Burkina Faso received either azithromycin or placebo in labour between October 2017 and May 2021. Because the intervention was given during labour, it should not affect the prevalence or development of LBW (17), hence we included the total cohort from both countries in these analyses.

### Setting

The PregnAnZI-2 trial was conducted at ten government health facilities in West Africa: two in The Gambia and eight in Burkina Faso(17). The two Gambian trial sites were in the Urban Western Health Region One: Serekunda Health Centre (SHC) and Bundung Maternal and Child Health Hospital (BMCHH). During the trial period, the sites collectively conducted over 7,000 deliveries/year. SHC provided Basic Emergency Obstetric and Newborn Care (BEmONC). All women requiring Comprehensive Emergency Obstetric and Newborn Care (CEmONC), including caesarean section, were referred to Kanifing General Hospital, a secondary-level facility located 3.4 kilometres (km) away. BMCHH provided CEmONC and rarely referred women to the tertiary level at the Edward Francis Small Teaching Hospital (EFSTH) in Banjul, located approximately 15 km away, when specialised interventions were required, or critical personnel were unavailable at BMCHH.

All eight sites in Burkina Faso were primary health facilities located in the rural districts of Nanoro and Yako. CEmONC was not available locally, and women requiring emergency caesarean section were referred to the Centre médical avec antenne chirurgicale (CMA) de Nanoro or CMA de Boussé, located maximum 25 km away. If a woman delivered at a facility different from the trial catchment facilities, this was recorded as “other,” and was predominantly a referral hospital where specialised expertise and CEmONC were available.

Both The Gambia and Burkina Faso have a Sahelian climate, typically hot and humid throughout the year with a long dry season (November to May) and a short rainy season (June to October) (18, 19). While most of the population in The Gambia were urban (61.9%) in 2019 (20), only 30.9% of Burkina Faso’s population lived in urban areas (21). Health facility deliveries were common in both countries in 2020, accounting for 84% of births in The Gambia (22), and up to 94% of births in Burkina Faso (23). The population of the study areas is representative of each country’s majority ethnic groups: Mandinka in The Gambia and Mossi in Burkina Faso (24, 25). The national LBW prevalence in The Gambia and Burkina Faso was 13.2% and 18.5% respectively in 2020, according to model estimates based on population-level data (3). In 2021, the stillbirth rate was 19 per 1000 total births in The Gambia and 21 per 1000 total births in Burkina Faso, with neonatal mortality rates of 25 per 1000 live births in both countries (26).

### Participants

Pregnant women aged ≥16 years who gave written informed consent during antenatal care visits were enrolled in the PregnAnZI-2 trial if they presented at a study health facility in active labour and verbally confirmed their willingness to participate (17). Women were excluded if they had known HIV infection; any other chronic or acute condition; planned caesarean section or known required referral for higher level obstetric care; known severe congenital malformation; confirmed intrauterine foetal death; allergy to macrolides; or use of drugs known to prolong the QT interval, e.g., chloroquine, erythromycin during the preceding two weeks (17). In this ancillary study, we included all offspring from recruited women who had documented birth weight, including stillbirths.

### Study procedures, data collection, and follow-up

Deliveries were managed by health facility staff, with research nurses and clinicians available to support with clinical care (17). Neonatal resuscitation was provided according to the Helping Babies Breathe protocol. We classified stillbirths as intrapartum stillbirths, as all enrolled women had a confirmed foetal heartbeat during labour. Stillbirths were identified by the attending facility staff on clinical grounds by detection of the absence of heart rate and breathing, with immediate documentation by the research nurse and clinician. The Apgar score was assigned at 1 and 5 minutes after delivery by the facility nurse or midwife who conducted or assisted delivery. Birth weight was measured for all neonates, including stillbirths, by a trained research nurse using calibrated electronic weighing scales (Seca 384/385) with a precision of 0.1 kg at birth or within 24 hours of delivery. Easily recognisable congenital malformations were identified during a detailed clinical examination by a research clinician or paediatrician between 4-24h after delivery and were classified post-hoc according to ICD-11 (27). A research nurse collected maternal sociodemographic and clinical data from the antenatal hand-held record. Active surveillance for outcome data was conducted 28 days after delivery by a research nurse in the participants’ homes in The Gambia and at a study health facility in Burkina Faso. A review was arranged by the study team if the mother or family was concerned about the neonate, with travel expenses and communication costs reimbursed, providing passive surveillance to 28 days. Any unwell neonate was reviewed by a study clinician and admitted or referred for appropriate neonatal care at either a study site or MRC The Gambia Clinical Services Department (Gambia only).

### Outcomes, variables of interest, and definitions

The primary outcome of this study was LBW. Based on the definition of LBW (<2.5kg), a binary variable was generated: 1 “LBW” (<2.5 Kg), and 0 “not LBW” (>=2.5 Kg). Neonates with a birth weight greater than 4kg (macrosomia) were excluded from the “not LBW” category as macrosomia is a distinct entity with specific risk factors and is not representative of normal fetal growth.

To explore factors associated with LBW, we adapted the Small Vulnerable Newborn (SVN) conceptual framework based on the biological plausibility and existing evidence for LBW risk factors due to foetal growth restriction, PTB, or both (Figure 1) (8, 9). In the adapted framework, we categorised variables in consecutive periods as pre-conception, conception, or pregnancy/antenatal, which were further categorised as maternal, placental, or foetal risk factors. Beyond biological, clinical, and epidemiological characteristics, the framework assumes the existence of contextual factors such as health care, environment/exposures, water sanitation hygiene, political, society/culture, and agriculture (8).

**Figure 1.**
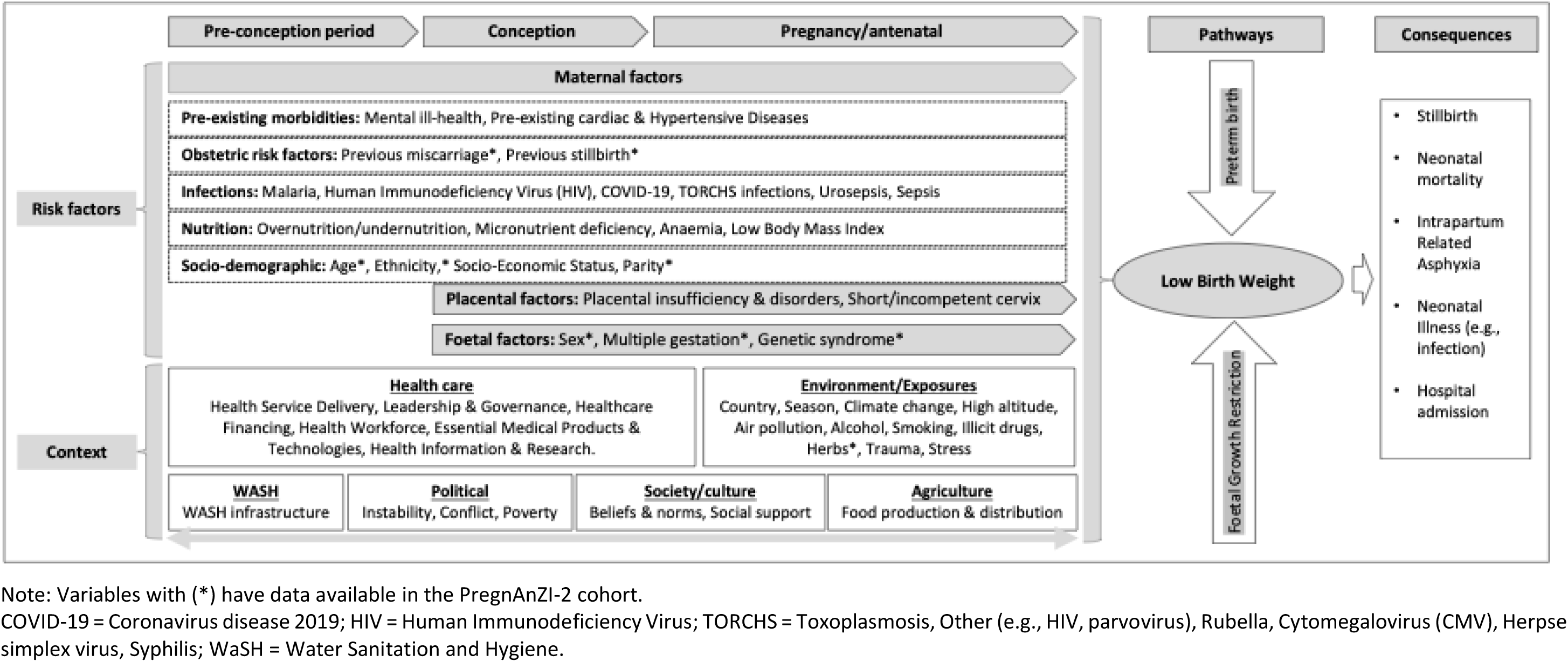
Conceptual framework for the development and short-term sequelae of low birth weight (LBW) in West Africa.

The short-term consequences of LBW that were considered included perinatal outcomes (stillbirth and intrapartum related asphyxia) and neonatal outcomes (hospital admission and neonatal mortality). Stillbirth was defined as the complete expulsion, or extraction, of a foetus following its in-utero demise at gestational age ≥22 weeks (27), including both fresh and macerated stillbirths. All stillbirths in this cohort are intrapartum as foetal demise occurred after the onset of labour and before delivery, known due to prospective documentation of fetal heartbeat during labour. An Apgar score of <7 at 1-minute after delivery was taken as a proxy for intrapartum-related asphyxia. Neonatal admission and neonatal death were defined as all hospitalisations and deaths that occurred during the 28-day follow-up period (28).

### Data management and statistical analysis

Data were collected electronically using encrypted mobile and computer devices at all study sites and uploaded to a study-specific Research Electronic Data Capture (REDCap) database. Data consistency checks and validation were conducted as per PregnAnZI-2 trial procedures (28).

Statistical analyses were performed in Stata/SE 18.5. The prevalence of LBW was calculated for the total cohort as well as stratified by birth outcome (live birth and stillbirth). Descriptive analyses were performed to determine the proportion of maternal and foetal explanatory (exposure) variables and adverse perinatal and neonatal outcomes as proportions of the total cohort (live births and stillbirths combined), stratified by LBW. For categorical variables, the baseline category was chosen to reflect either a “normal pregnancy and delivery” (e.g., non-extremes of parity) or the most prevalent grouping (e.g., ethnicity). A univariate logistic regression model was then fitted, and each exposure variable was selected according to the conceptual framework (Figure 1) if data from the PregnAnZI-2 database were available. An adjusted logistic regression model was developed for each exposure variable with a p-value <0.2 on univariate analysis. To avoid over-adjusting for multiple variables on the same causal pathway (29), each model included only variables occurring before or at the same time as the exposure variable of interest (Figure 1). As ethnicity and country were co-linear with no overlap, we adjusted only for ethnicity to provide more granular insights to risk factors for LBW. Statistical significance for the adjusted analysis was pre-determined at *p<0.05.* Exposure variables with more than 10% of missing data were excluded from the risk factor analysis, with available case analysis used for variables with less than 10% of missing data.

## RESULTS

### Participant Overview

11,980 women and their 12027 offspring were included (Figure 2). Overall, 9.6% (1159/12027) of the newborns were LBW. The proportion of LBW was higher among stillbirths (26.2%, 22/84) compared to live births (9.5%, 1137/11943, p<0.001) (Figure 2). The prevalence of LBW was lower in The Gambia compared to Burkina Faso for the total cohort (8.2% versus 11.4%, p<0.001)(Table 1).

**Figure 2.**
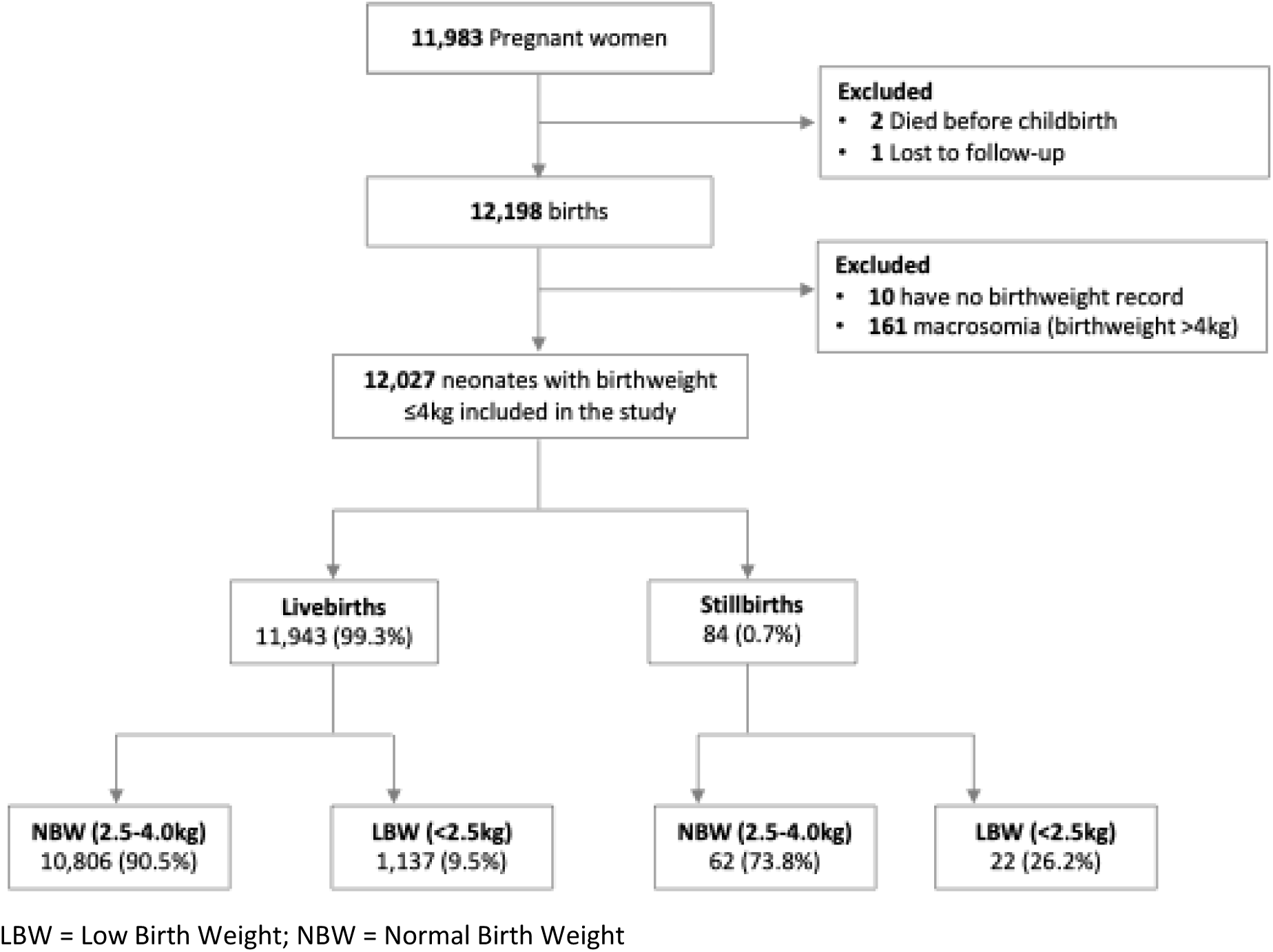
Overview of study participants and perinatal outcomes from a cohort of West African pregnant women.

**Table 1.**
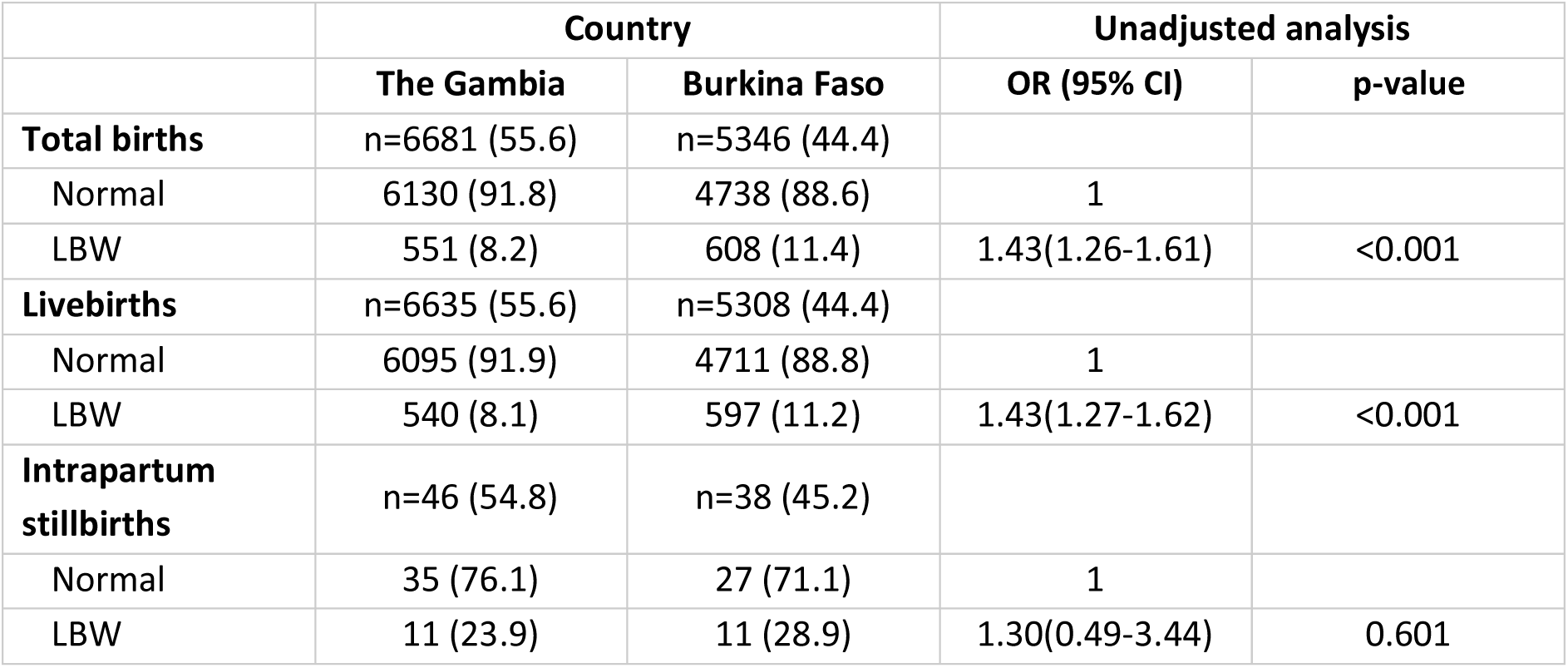
Low birth weight prevalence by perinatal outcome, stratified by country.

|  | Country |  | Unadjusted analysis |  |
| --- | --- | --- | --- | --- |
|  | The Gambia | Burkina Faso | OR (95% CI) | p-value |
| **Total births** | n=6681 (55.6) | n=5346 (44.4) |  |  |
| Normal | 6130 (91.8) | 4738 (88.6) | 1 |  |
| LBW | 551 (8.2) | 608 (11.4) | 1.43(1.26-1.61) | <0.001 |
| **Livebirths** | n=6635 (55.6) | n=5308 (44.4) |  |  |
| Normal | 6095 (91.9) | 4711 (88.8) | 1 |  |
| LBW | 540 (8.1) | 597 (11.2) | 1.43(1.27-1.62) | <0.001 |
| **Intrapartum stillbirths** | n=46 (54.8) | n=38 (45.2) |  |  |
| Normal | 35 (76.1) | 27 (71.1) | 1 |  |
| LBW | 11 (23.9) | 11 (28.9) | 1.30(0.49-3.44) | 0.601 |

### Risk factors associated with LBW

Pre-conception factors associated with LBW included maternal ethnicity, parity, and previous history of stillbirth. Gourounsi ethnicity was associated with an increased odds of LBW compared to Mossi ethnicity (aOR:1.24, 95%CI 0.90-1.70, p<0.001). Primiparous mothers had higher odds of delivering a LBW neonate compared to those with a parity of 2 (aOR:1.63, 95%CI 1.33-1.99, p<0.001). A maternal history of stillbirth was also associated with LBW compared to women without a previous stillbirth (aOR:1.88, 95%CI 1.35-2.62, p<0.001). Among foetal factors, twins (aOR 22.96, 95%CI 18.51-28.48, p<0.001) and female neonates (aOR 1.58, 95%CI 1.38-1.80, p<0.001) were associated with LBW (Table 2).

**Table 2.** Risk factors for LBW following relatively healthy pregnancy in The Gambia and Burkina Faso.

|  | Total<br>n | LBW<br>n (%) | Normal birth<br>weight<br>n (%) | Unadjusted analysis |  | Adjusted analysis |  |
| --- | --- | --- | --- | --- | --- | --- | --- |
|  |  |  |  | cOR<br>(95%CI) | p-value | aOR<br>(95%CI) | p-value |
|  | **n=12,027** | **n=1,159** | **n=10,868** |  |  |  |  |
| **Maternal pre-conception factors** |  |  |  |  |  |  |  |
| **Country of residence** |  |  |  |  |  |  |  |
| The Gambia | 6681 | 551 (8.3) | 6130 (91.8) | 1 | <0.001 |  |  |
| Burkina Faso | 5346 | 608 (11.4) | 4738 (88.6) | 1.43(1.26-1.61) |  |  |  |
| **Maternal age (yrs)** |  |  |  |  |  |  |  |
| Median (IQR) | 26(22-31) |  |  |  |  |  |  |
| 20 – 35 | 9407 | 844 (9.0) | 8559 (91.0) | 1 | <0.001 | 1<sup>a</sup> | 0.067 |
| 16 – 19 | 1524 | 232 (15.2) | 1291 (84.8) | 1.82(1.56-2.13) |  | 1.20 (0.99-1.47) |  |
| >35 | 1101 | 83 (7.5) | 1018 (92.6) | 0.82(0.65-1.05) |  | 0.96(0.75-1.23) |  |
| **Maternal ethnicity** |  |  |  |  |  |  |  |
| Mossi | 4924 | 551 (11.2) | 4373 (88.8) | 1 | <0.001 | 1<sup>a</sup> | <0.001 |
| Gourounsi | 346 | 48 (13.9) | 298 (86.1) | 1.28(0.93-1.76) |  | 1.24(0.90-1.70) |  |
| Other BF | 76 | 9 (11.8) | 67 (88.2) | 1.07(0.53-2.15) |  | 1.08(0.53-2.18) |  |
| Mandinka | 2671 | 226 (8.5) | 2445 (91.5) | 0.73(0.62-0.86) |  | 0.73(0.62-0.86) |  |
| Fula | 1200 | 99 (8.3) | 1101 (91.7) | 0.71(0.57-0.89) |  | 0.67(0.53-0.84) |  |
| Wolof | 1024 | 102 (10.0) | 922 (90.0) | 0.88(0.70- 1.10) |  | 0.87(0.69-1.09) |  |
| Jola | 860 | 52 (6.1) | 808 (93.9) | 0.51(0.38-0.69) |  | 0.50(0.37-0.68) |  |
| Other Gambia | 926 | 72 (7.8) | 854 (92.2) | 0.67(0.52-0.86) |  | 0.66(0.51-0.85) |  |
| **Parity** |  |  |  |  |  |  |  |
| Para 2 | 2222 | 192 (8.6) | 2030 (91.4) | 1 | <0.001 | 1<sup>a</sup> | <0.001 |
| Primiparous | 2738 | 382 (14.0) | 2356 (86.0) | 1.71(1.43-2.06) |  | 1.63(1.33-1.99) |  |
| Para 3 | 1947 | 174 (8.9) | 1773 (91.1) | 1.04(0.84-1.29) |  | 1.05(0.84-1.30) |  |
| Para ≥4 | 5120 | 411 (8.0) | 4709 (92.0) | 0.92(0.77-1.10) |  | 0.89(0.73-1.07) |  |
| **Previous miscarriage** |  |  |  |  |  |  |  |
| No | 10907 | 1052 (9.7) | 9855 (90.3) | 1 | 0.921 |  |  |
| Yes | 1120 | 107 (9.6) | 1013 (90.4) | 0.99(0.80-1.22) |  |  |  |
| **Previous stillbirth** |  |  |  |  |  |  |  |
| No | 11735 | 1115 (9.5) | 10620 (90.5) | 1 | 0.002 | 1<sup>a</sup> | <0.001 |
| Yes | 292 | 44 (15.1) | 248 (84.9) | 1.69(1.22-2.34) |  | 1.88(1.35-2.62) |  |
| **Maternal pregnancy factors** |  |  |  |  |  |  |  |
| **Traditional medicine use** |  |  |  |  |  |  |  |
| No | 10854 | 1069 (9.9) | 9785 (90.1) | 1 | 0.017 | 1<sup>b</sup> | 0.164 |
| Yes | 1173 | 90 (7.7) | 1083 (92.3) | 0.76(0.61-0.95) |  | 0.84(0.65-1.08) |  |
| **Foetal conception and pregnancy factors** |  |  |  |  |  |  |  |
| **Sex of newborn** |  |  |  |  |  |  |  |
| Male | 6179 | 490 (7.9) | 5689 (92.1) | 1 | <0.001 | 1<sup>c</sup> | <0.001 |
| Female | 5848 | 669 (11.4) | 5179 (88.6) | 1.50(1.33-1.70) |  | 1.58(1.38-1.80) |  |
| **Plurality of birth** |  |  |  |  |  |  |  |
| Singleton | 11595 | 901 (7.8) | 10694 (92.2) | 1 | <0.001 | 1<sup>c</sup> | <0.001 |
| Twin | 432 | 258 (59.7) | 174 (40.3) | 17.60(14.35-21.58) |  | 22.96(18.51-28.48) |  |
| **Easily recognisable congenital malformations** |  |  |  |  |  |  |  |
| No | 11813 | 1137 (9.6) | 10,676 (90.4) | 1 | 0.748 |  |  |
| Yes | 214 | 22 (10.3) | 192 (89.7) | 1.08(0.69-1.68) |  |  |  |

a) Maternal Pre-conception Factors - adjusted for maternal age, ethnicity, parity, and previous stillbirth.
b) Maternal Conception and Pregnancy Factors - adjusted for maternal age, ethnicity, parity, and previous stillbirth.
c) Foetal Conception and Pregnancy Factors - adjusted for maternal age, ethnicity, parity, previous stillbirth, sex, and plurality (twins).
cOR = Crude odds ratio; aOR = Adjusted odds ratio; IQR = Interquartile range; BF = Burkina Faso.

### Short-term adverse outcomes associated with LBW

Being born LBW increased the risk of adverse outcomes including neonatal mortality (OR 4.35, 95% CI 3.25-5.81, p<0.001), intrapartum stillbirth (OR 3.37, 95% CI 2.07-5.51, p<0.001), hospital admission (OR 2.17, 95% CI 1.70-2.77, p<0.001), and intrapartum related asphyxia (OR 2.04, 95% CI 1.59-2.62, p<0.001)(Table 3). LBW neonates contributed disproportionately to these adverse outcomes, accounting for 30.7% (69/225) of neonatal deaths, 26.2% (22/84) of intrapartum stillbirths, 18.2% (85/467) of admissions, and 17.4% (80/461) of neonates born with low 1-minute apgar score (Table 3).

**Table 3.** Adverse short-term outcomes associated with low birth weight following low-risk pregnancy in The Gambia and Burkina Faso.

|  | Total<br>n(%) | LBW<br>n (%) | Normal birth weight<br>n (%) | Unadjusted analysis |  |
| --- | --- | --- | --- | --- | --- |
|  |  |  |  | OR (95%CI) | p-value |
| **Intrapartum stillbirth** | **N=12027** | **N=1,159** | **N=10,868** |  |  |
| No | 11943 (99.3) | 1137 (9.5) | 10806 (90.5) | 1 | <0.001 |
| Yes | 84 (0.7) | 22 (26.2) | 62 (73.8) | 3.37(2.07-5.51) |  |
| **1 min Apgar score <7** | **n=12011<sup>a</sup>** |  |  |  |  |
| No | 11550 (96.2) | 1076 (9.3) | 10474 (90.7) | 1 | <0.001 |
| Yes | 461 (3.8) | 80 (17.4) | 381 (82.6) | 2.04(1.59-2.62) |  |
| **Neonatal admissions by 28 days** | **n=12027** |  |  |  |  |
| No | 11560 (96.1) | 1074 (9.3) | 10486 (90.7) | 1 | <0.001 |
| Yes | 467 (3.9) | 85 (18.2) | 382 (81.8) | 2.17(1.70-2.77) |  |
| **Neonatal mortality by 28 days** | **n=12027** |  |  |  |  |
| No | 11802 (98.1) | 1090 (9.2) | 10712 (90.8) | 1 | <0.001 |
| Yes | 225 (1.9) | 69 (30.7) | 156 (69.3) | 4.35(3.25-5.81) |  |

a) N=16 had missing data for 1-minute apgar score

## DISCUSSION

Although LBW affected less than 10% of births in this study, these infants accounted for a disproportionately high burden of adverse outcomes. Almost 1 in 4 (24%) of deaths and 1 in 5 (19%) of intrapartum stillbirths were attributable to LBW. Similarly, approximately 1 in 10 IRAs (9%) and neonatal hospitalisations (10%), respectively, are LBW. We also identified several non-modifiable yet easily identifiable maternal (primiparity and history of previous stillbirth) and foetal (twin and female sex) risk factors for LBW.

The overall prevalence of LBW in our cohort was 9.6% and was higher in Burkina Faso (11.4%) than in The Gambia (8.3%). This intra-regional difference may reflect the rural setting in Burkina Faso, where risk factors for LBW, such as malaria, are more prevalent (30). Our findings are consistent with previous estimates from The Gambia (10.5%)(31), Burkina Faso (11.0%)(30), Ghana (9.7%)(32) and a meta-analysis of DHS data from 35 countries in Sub-Saharan Africa (SSA) reporting a LBW prevalence of 9.7% (33). While under-reporting is common in most population-based studies due to poor recall and exclusion of stillbirths, our exclusion of high-risk pregnancies likely resulted in a conservative estimate, suggesting that the population-level burden may be even higher. Studies from Ethiopia have reported substantially higher rates of LBW, ranging from 16-34%, likely due to poor antenatal care, with up to 48% of mothers in one study receiving no antenatal visits (34–36).

The increased risk of adverse outcomes such as intrapartum stillbirth, intrapartum related asphyxia, and neonatal mortality among neonates with LBW mirrors findings from similar settings (34) and underlines that LBW neonates are a highly vulnerable group. The increased risk of poor outcome among LBW neonates in our cohort is reflected by them accounting for 23.7% of neonatal deaths. LBW was also a main contributor to intrapartum stillbirth. Therefore, formulating policies to identify and manage LBW pregnancies and those at risk of premature delivery will be critical for improving pregnancy and postnatal outcomes. These policies should include early screening for LBW using accessible tools such as simple and structured obstetric history taking, fundal height measurements, or routine ultrasounds (37, 38). Proven antenatal interventions such as multiple micronutrient supplements, screening and treatment for asymptomatic bacteriuria, balanced energy and protein supplements, low-dose aspirin, zinc supplements, and calcium supplements should also be promoted during antenatal care (13).

We identified several maternal and foetal factors associated with LBW. Although most of these factors are non-modifiable, they can be detected early to help identify pregnancies at highest risk of LBW. Among maternal preconception factors shown in the conceptual framework (Figure 1), primiparity and a history of stillbirth were the most prominent. Primiparity is a well-established risk factor for LBW in SSA countries, including The Gambia (31), Burkina Faso, and Ghana (39). This association is possibly driven by inadequate physiologic adaptations (e.g., reduced uteroplacental blood flow) (40) and anatomical limitations on uterine capacity during the first pregnancy (41). These findings reinforce the importance of strengthening preconception care and prioritising targeted support for first-time pregnant women. Similarly, women with a history of stillbirth should be considered a high-risk group (42) and provided with enhanced antenatal and intrapartum monitoring. Potential underlying causes such as sickle cell disease, cervical insufficiency, recurrent infections, or psychological stressors like domestic violence or occupational stress, should be actively investigated during pregnancy, as illustrated in Figure 1 above. Health care providers should move beyond recording previous stillbirths to understanding the underlying causes and tailoring care accordingly. Foetal factors associated with LBW in our study included twin gestation and female sex, consistent with previous research across SSA (33). In settings with poor healthcare (Figure 1) where access to reliable and affordable antenatal ultrasound remains limited, prioritising scans for high-risk twin pregnancies to monitor growth and plan safe delivery could optimise the use of scarce diagnostic resources while improving foetal surveillance and birth outcomes.

The main strength of our study is its contribution to filling a critical data gap on LBW in relation to intrapartum stillbirth. The quality of data is also important, as this analysis was embedded in a randomised clinical trial, ensuring a robust and standardised data collection, including accurate measurement of birth weight data. However, several limitations should also be acknowledged. First, the exclusion of pregnant women with acute and chronic conditions may have underestimated the prevalence of LBW, limiting the generalisability of our findings. Second, the absence of gestational age data prevented us from distinguishing between LBW phenotypes (e.g., preterm LBW, growth-restricted LBW, or both), which limits our ability to explore distinct causal pathways (43). Third, the use of secondary data constrained the scope of the risk factors analysis, with key variables such as malaria and anaemia in pregnancy unavailable. The need for more comprehensive investigation of risk factors associated with LBW in West Africa warrants prospective cohort studies to identify potentially modifiable risk factors which could strengthen antenatal care, optimise foetal nutrition and growth, prevent PTB and facilitate safe intrapartum management. Lastly, although the PregnAnZI-2 intervention was given during labour and should not have affected development of LBW or PTB, we do not know if azithromycin had a birth weight dependent effect on the risk of adverse perinatal and neonatal outcomes.

## CONCLUSION

Our findings highlight that LBW is a major contributor to intrapartum stillbirth, IRA, neonatal admission, and neonatal mortality in West Africa. The identification of easily recognisable maternal and foetal risk factors such as primiparity, history of stillbirth and twin gestation, among others, offers an opportunity to strengthen antenatal risk stratification and intrapartum management. In resource-limited settings, prioritising targeted interventions for pregnancies high-risk for LBW could substantially reduce adverse outcomes, accelerating progress toward achieving Sustainable Development Goal 3.2.

## DECLARATIONS

### Ethical approval & consent to participate

The PregnAnZI-2 trial was approved by The Gambia Government/Medical Research Council Unit, The Gambia (MRCG) Joint Ethics Committee, the Comité d’Ethique pour la Recherche en Santé (CERS) and the Ministry of Health of Burkina Faso, and the LSHTM Ethics Committee. This included approval for further research utilising trial data. All women provided written informed consent for trial participation during antenatal care visits and were free to withdraw at any time.

### Availability of data & materials

Data may be obtained from a third party and is not publicly available. The clinical data have been collected following the provision of informed consent, under the prerequisite of strict participant confidentiality. Qualified researchers may request access through the Gambia Government/MRC Joint Ethics Committee. The review process and release of data will be facilitated by the MRC Unit The Gambia (http://www.mrc.gm/) through the Head of Governance at MRCG at LSHTM. Access will not be unduly restricted.

### Authors’ contributions

AR and UDA conceived and designed the PregnAnZI-2 trial. BC designed this study with input from AR, UDA, HB, and CB. BC, JB, and PregnAnZI-2 field teams in The Gambia and Burkina Faso coordinated data collection. BC performed the analysis with full access to the data and input from CB. BC drafted the initial manuscript with inputs from HB, AR, CB, and UDA. All authors contributed to the final version. AR provided oversight of the work as the guarantor and accepts full responsibility for the finished work and controlled the decision to publish.

## Acknowledgments

We thank the project manager, Asheme Mahmoud, and the senior clinical trial assistant, Bakary Fatty (who subsequently acted as the project officer), Maxine Haffner, and all the field, data management, and laboratory teams for providing support towards the conduct of the main trial. We also thank the leadership board and staff health facilities in The Gambia and Burkina Faso for their support. We are grateful to all the mothers and their neonates whose data was used in this study following their participation in the PregnAnZI-2 trial.

## REFERENCES

1. WHO recommendations for care of the preterm or low birth weight infant. Geneva: World Health Organization; 2022.

2. Okwaraji YB, Suárez-Idueta L, Ohuma EO, Bradley E, Yargawa J, Pingray V, et al. Stillbirth risk by fetal size among 126.5 million births in 15 countries from 2000 to 2020: A fetuses-at-risk approach. Bjog. 2024.

3. World Health Organization, United Nations Children’s F. Low birthweight estimates: levels and trends 2000-2020. New York; 2023.

4. Christian P, Lee SE, Donahue Angel M, Adair LS, Arifeen SE, Ashorn P, et al. Risk of childhood undernutrition related to small-for-gestational age and preterm birth in low- and middle-income countries. Int J Epidemiol. 2013;42(5):1340–55.

5. Breslau N, DelDotto JE, Brown GG, Kumar S, Ezhuthachan S, Hufnagle KG, et al. A gradient relationship between low birth weight and IQ at age 6 years. Arch Pediatr Adolesc Med. 1994;148(4):377–83.

6. Jornayvaz FR, Vollenweider P, Bochud M, Mooser V, Waeber G, Marques-Vidal P. Low birth weight leads to obesity, diabetes and increased leptin levels in adults: the CoLaus study. Cardiovasc Diabetol. 2016;15:73.

7. Jamaluddine Z, Sharara E, Helou V, El Rashidi N, Safadi G, El-Helou N, et al. Effects of size at birth on health, growth and developmental outcomes in children up to age 18: an umbrella review. Archives of Disease in Childhood. 2023;108(12):956–69.

8. Ashorn P, Ashorn U, Muthiani Y, Aboubaker S, Askari S, Bahl R, et al. Small vulnerable newborns&#x2014;big potential for impact. The Lancet. 2023;401(10389):1692-706.

9. Hunter PJ, Awoyemi T, Ayede AI, Chico RM, David AL, Dewey KG, et al. Biological and pathological mechanisms leading to the birth of a small vulnerable newborn. The Lancet. 2023;401(10389):1720–32.

10. Cutland CL, Lackritz EM, Mallett-Moore T, Bardají A, Chandrasekaran R, Lahariya C, et al. Low birth weight: Case definition & guidelines for data collection, analysis, and presentation of maternal immunization safety data. Vaccine. 2017;35(48, Part A):6492-500.

11. Damhuis SE, Ganzevoort W, Gordijn SJ. Abnormal Fetal Growth: Small for Gestational Age, Fetal Growth Restriction, Large for Gestational Age: Definitions and Epidemiology. Obstet Gynecol Clin North Am. 2021;48(2):267–79.

12. Khandre V, Potdar J, Keerti A. Preterm Birth: An Overview. Cureus. 2022;14(12):e33006.

13. Hofmeyr GJ, Black RE, Rogozińska E, Heuer A, Walker N, Ashorn P, et al. Evidence-based antenatal interventions to reduce the incidence of small vulnerable newborns and their associated poor outcomes. The Lancet. 2023;401(10389):1733–44.

14. WHO. Global nutrition targets 2025: low birth weight policy brief (WHO/NMH/NHD/14.5. Geneva: World Health Organization. 2014.

15. Okwaraji YB, Krasevec J, Bradley E, Conkle J, Stevens GA, Gatica-Domínguez G, et al. National, regional, and global estimates of low birthweight in 2020, with trends from 2000: a systematic analysis. The Lancet. 2024;403(10431):1071–80.

16. Blencowe H CS, Oestergaard M, Chou D, Moller AB, Narwal R, Adler A, Garcia CV, Rohde S, Say L, Lawn JE. National, regional and worldwide estimates of preterm birth rates in the year 2015 with time trends since 1990 for selected countries: a systematic analysis and implications. 2015.

17. Roca A, Camara B, Bognini JD, Nakakana UN, Somé AM, Beloum N, et al. Effect of Intrapartum Azithromycin vs Placebo on Neonatal Sepsis and Death: A Randomized Clinical Trial. JAMA. 2023;329(9):716–24.

18. Government of The Gambia Ministry of Environment CCaNR. THE GAMBIÀS LONG-TERM CLIMATE-NEUTRAL DEVELOPMENT STRATEGY 2050. 2022.

19. Group WB. G5 Sahel Region Country Climate and Development Report. © Washington, DC: World Bank; 2022.

20. Pantaleo C. The Gambia: Consultancy for Urban Targeting for The Gambia Social Registry. 2021.

21. WCARCO U. Burkina Faso: Monographic Study on Demography, Peace, And Security in The Sahel. 2020.

22. Gambia Bureau of Statistics - GBoS, ICF. The Gambia Demographic and Health Survey 2019-20. Banjul, The Gambia: GBoS/ICF; 2021.

23. Institut National de la Statistique et de la Démographie, The DHS Program. Burkina Faso Enquête Démographique et de Santé 2021. Burkina Faso et Rockville, Maryland, USA: INSD et ICF; 2023.

24. Gambia Bureau of Statistics - GBoS. Population and Housing Census Spatial Distribution. 2013.

25. Central Census Bureau (Burkina Faso). Burkina Faso Population and Housing Census. Ministry of Economy, Finance and Planning 2019.

26. United Nations Inter-agency Group for Child Mortality Estimation (UN IGME). Never Forgotten: The situation of stillbirth around the globe, United Nations Children’s Fund,. New York; 2023.

27. World Health Organization. ICD-11 International Classification of Diseases 11th Revision. The Gold Standard for Diagnostic Health Information 2019.

28. Camara B, Bognini JD, Nakakana UN, Some AM, Jagne I, Tahita MC, et al. Pre-delivery administration of azithromycin to prevent neonatal sepsis and death: a phase iii double-blind randomized clinical trial (PregnAnZI-2 trial). 2022. 2022;9(1):11.

29. Westreich D, Greenland S. The table 2 fallacy: presenting and interpreting confounder and modifier coefficients. Am J Epidemiol. 2013;177(4):292–8.

30. Lingani M, Zango SH, Valéa I, Somé G, Sanou M, Samadoulougou SO, et al. Low birth weight and its associated risk factors in a rural health district of Burkina Faso: a cross sectional study. BMC Pregnancy and Childbirth. 2022;22(1):228.

31. Jammeh A, Sundby J, Vangen S. Maternal and obstetric risk factors for low birth weight and preterm birth in rural Gambia: a hospital-based study of 1579 deliveries. Open Journal of Obstetrics and Gynecology. 2011;1, 94–103.

32. Agbozo F, Abubakari A, Der J, Jahn A. Prevalence of low birth weight, macrosomia and stillbirth and their relationship to associated maternal risk factors in Hohoe Municipality, Ghana. Midwifery. 2016;40:200–6.

33. Tessema ZT, Tamirat KS, Teshale AB, Tesema GA. Prevalence of low birth weight and its associated factor at birth in Sub-Saharan Africa: A generalized linear mixed model. PLOS ONE. 2021;16(3):e0248417.

34. Desta M, Tadese M, Kassie B, Gedefaw M. Determinants and adverse perinatal outcomes of low birth weight newborns delivered in Hawassa University Comprehensive Specialized Hospital, Ethiopia: a cohort study. BMC Res Notes. 2019;12(1):118.

35. Mehare T, Sharew Y. Prevalence and Associated Factors of Low Birth Weight among Term Newborns in Dilla Town, Southern Ethiopia. Int J Pediatr. 2020;2020:8394578.

36. Endalamaw A, Engeda EH, Ekubagewargies DT, Belay GM, Tefera MA. Low birth weight and its associated factors in Ethiopia: a systematic review and meta-analysis. Ital J Pediatr. 2018;44(1):141.

37. Papageorghiou AT, Ohuma EO, Gravett MG, Hirst J, da Silveira MF, Lambert A, et al. International standards for symphysis-fundal height based on serial measurements from the Fetal Growth Longitudinal Study of the INTERGROWTH-21st Project: prospective cohort study in eight countries. Bmj. 2016;355:i5662.

38. Morse K, Williams A, Gardosi J. Fetal growth screening by fundal height measurement. Best Pract Res Clin Obstet Gynaecol. 2009;23(6):809–18.

39. He Z, Bishwajit G, Yaya S, Cheng Z, Zou D, Zhou Y. Prevalence of low birth weight and its association with maternal body weight status in selected countries in Africa: a cross-sectional study. BMJ Open. 2018;8(8):e020410.

40. Clapp JF, 3rd, Capeless E. Cardiovascular function before, during, and after the first and subsequent pregnancies. Am J Cardiol. 1997;80(11):1469–73.

41. Sørnes T, Bakke T. Uterine size, parity and umbilical cord length. Acta Obstet Gynecol Scand. 1989;68(5):439–41.

42. Ladhani NNN, Fockler ME, Stephens L, Barrett JFR, Heazell AEP. No. 369-Management of Pregnancy Subsequent to Stillbirth. Journal of Obstetrics and Gynaecology Canada. 2018;40(12):1669–83.

43. Lawn JE, Ohuma EO, Bradley E, Idueta LS, Hazel E, Okwaraji YB, et al. Small babies, big risks: global estimates of prevalence and mortality for vulnerable newborns to accelerate change and improve counting. The Lancet. 2023;401(10389):1707–19.

